# Aging-dependent Change in Th17 and Cytokine Response in Multiple Sclerosis

**DOI:** 10.1101/2024.03.17.24304425

**Authors:** Wen Zhu, Shankar Revu, Chenyi Chen, Megan Dahl, Archana Ramkumar, Conor Kelly, Mandy J McGeachy, Zongqi Xia

**Affiliations:** Department of Neurology, University of Pittsburgh, Pittsburgh, PA, USA; Department of Medicine, University of Pittsburgh, Pittsburgh, PA, USA; Department of Microbiology and Immunology, College of Veterinary Medicine, Cornell University, Ithaca, NY, USA

**Keywords:** Multiple sclerosis, myelin basic protein, Th17, aging, relapse, inflammatory disease activity

## Abstract

**Background:** Multiple sclerosis (MS) is a chronic autoimmune disease damaging the central nervous system. Diminished inflammatory disease activity (DA) as people with MS (pwMS) age motivated randomized clinical trials assessing disease-modifying therapy (DMT) discontinuation in older pwMS given the concern for risks outweighing benefits. This study aims to examine whether peripheral production of Myelin Basic Protein (MBP)-driven cytokine responses mediate the aging-associated decline in MS inflammatory DA.

**Methods:** We included the clinical data of 669 adult pwMS between 2017 and 2022 who enrolled in a clinic-based prospective cohort. From a subset of 80 participants, we isolated fresh peripheral blood mononuclear cells (PBMCs) and cultured with 50μg/ml of MBP (or heat-killed Candida) for 24 hours. We assayed cell culture supernatants for interleukin 17 (IL-17) and interferon gamma (IFN-γ) using Enzyme-Linked Immunosorbent Assay and a subset of the supernatant samples using a commercial human cytokine/chemokine array. We examined the associations between age and annualized relapse rate (ARR) as well as between age and MBP-stimulated cytokine production (by cultured PBMC) using covariate-adjusted linear regressions. We performed mediation analyses to determine the extent to which MBP-driven cytokine response drives the association between age and ARR.

**Results:** Among 669 pwMS (mean age 51.7±12.7 years, 80.7% women, 89.4% non-Hispanic White), ARR declined with age (β=-0.003, p<0.001). Among the subgroup of 80 pwMS whose cultured PBMCs underwent ex vivo MBP stimulation, IL-17 production declined with age in women (β=-0.27, p=0.04) but not men (β=-0.1, p=0.73). MBP-driven IL-17 response partially mediated the association between older age and lower ARR (24.7% in women, 15.3% in men). In exploratory analyses, older pwMS (≥50 years) had marginally lower (IL-4, MCP-2, MCP-3, PDGF-AA, PDGF-AB/BB) and higher (Fractalkine, MDC) concentrations of several cytokines than younger pwMS (<50 years), while certain cytokines (MCP-2, MDC) mediated whereas others negated the effect of age on ARR.

**Conclusion:** Diminished peripheral IL-17 response as a potential biological mechanism underlying the aging-dependent decline in MS inflammatory DA warrants further investigation.

## Introduction

Multiple sclerosis (MS) is a chronic autoimmune disease that causes inflammatory demyelination and progressive neurodegeneration in the central nervous system (CNS). Aging correlates with MS disease progression.[1,2] In people with MS (pwMS), 45 years of age seem to mark an inflection point as the relapse rate declines while disability accrual increases.[3] Both the inflammatory disease activity and the potential for a full recovery after a relapse diminish with increasing age,[4] potentially due to the diminished CNS “reserve” and the decreasing capacity to restore function with aging.[2] These considerations have motivated randomized clinical trials testing the safety of immune modulating therapy discontinuation in older pwMS with stable inflammatory disease activity given the concern for risks outweighing benefits.[5,6]

Interleukin-17 (IL-17) is a cytokine that promotes blood-brain barrier permeability, pro-inflammatory signaling in astrocytes, neuronal hyperexcitability, and depletion of oligodendrocyte precursors necessary for repairing the damaged CNS.[7,8] Consistent with its role in driving autoimmunity, IL-17 enhances B cell activation and autoantibody production in models of MS,[9,10] while increased IL-17 expression is found at sites of acute and chronic inflammation in pwMS.[11] Myelin-reactive CD4+ T helper (Th) cells, particularly Th17 cells that produce IL-17, are crucial in MS pathogenesis. Depletion of Th17 cells in a murine model of experimental autoimmune demyelination protects against the induction of MS-like pathology and phenotype.[12] Th17 cells are more abundant in the blood and cerebrospinal fluid of pwMS than healthy controls, and they further expand during active MS relapses.[13–16] Th17 cells encompass subpopulations that differentially produce IL-17 alone or in combination with other proinflammatory cytokines such as interferon-gamma (IFNγ) with the latter “Multi-functional” Th17 cells considered as more pathogenic. Overall Th17 cell frequency is higher in both people with relapsing-remitting MS (RRMS) and secondary-progressive MS (SPMS) than in healthy controls, while double-positive IL-17^+^ IFNγ^+^ Th17 cells are higher in the inflammation-predominant RRMS than the neurodegeneration-predominant SPMS.[17]

Immunosenescence and inflammaging potentially explain the aging-associated decline in MS disease activity. Immunosenescence describes dysregulated adaptive and innate immune responses that increase the risk of infection and malignancy as people age, while inflammaging refers to the low-grade chronic inflammation throughout the aging process that produces a broad range of tissue damages and chronic conditions. The former may contribute to the decline in acute inflammatory demyelination events while the latter may contribute to the disability accrual and disease progression with aging.[18] Among the cells crucial in MS pathogenesis and progression, Th17 cells also play a significant role in mediating inflammaging.[19,20] In a study of 122 pwMS, aging and Th17-dominant immune response profile (a higher Th17/Treg ratio after positively weighting IL-6 and IL-17 for Th17 and negatively weighting IL-10 for Treg) were independently associated with MS disability.[21]

Few studies to date have examined immune drivers of aging-related decline in MS relapse rate, particularly the role of Th17 cells. Myelin basic protein (MBP) is an abundant protein in the CNS myelin and a putative MS auto-antigen, and cytokines released from MBP-stimulated peripheral immune cells isolated from pwMS represent an experimental paradigm to investigate MS-relevant mechanisms.[22–30] In this study, we examined whether MBP-induced Th17/IL-17 and other cytokine responses mediate the aging-associated decline in MS relapse.

## Methods

### Study Design, Participants and Samples

This cross-sectional study included 669 participants (**primary cohort**) who enrolled in a clinic-based prospective cohort study (Prospective Investigation of Multiple Sclerosis in the Three Rivers Region, PROMOTE; Pittsburgh, PA) during 2017-2022 (Figure 1).[31–42] The cohort enrollment criteria included adults 18 years or older with a neurologist-confirmed diagnosis of MS according to the 2017 McDonald criteria. Given the clinical outcome of interest is relapse event indicative of inflammatory disease activity, we included pwMS of relapsing remitting (RRMS) and secondary progressive (SPMS) type and excluded primary progressive type (PPMS), which is not characterized by relapse events. To collect blood sample for MBP-stimulation assays, we randomly selected a subset of 80 participants (**Th17 cohort**) to achieve a 1:2 men to women ratio and relatively balanced distribution across age groups from 25 to 75 years old. Participants donated venous blood samples for research during routine clinical appointments. The blood samples were immediately processed for the downstream experiments.

**Figure 1.**
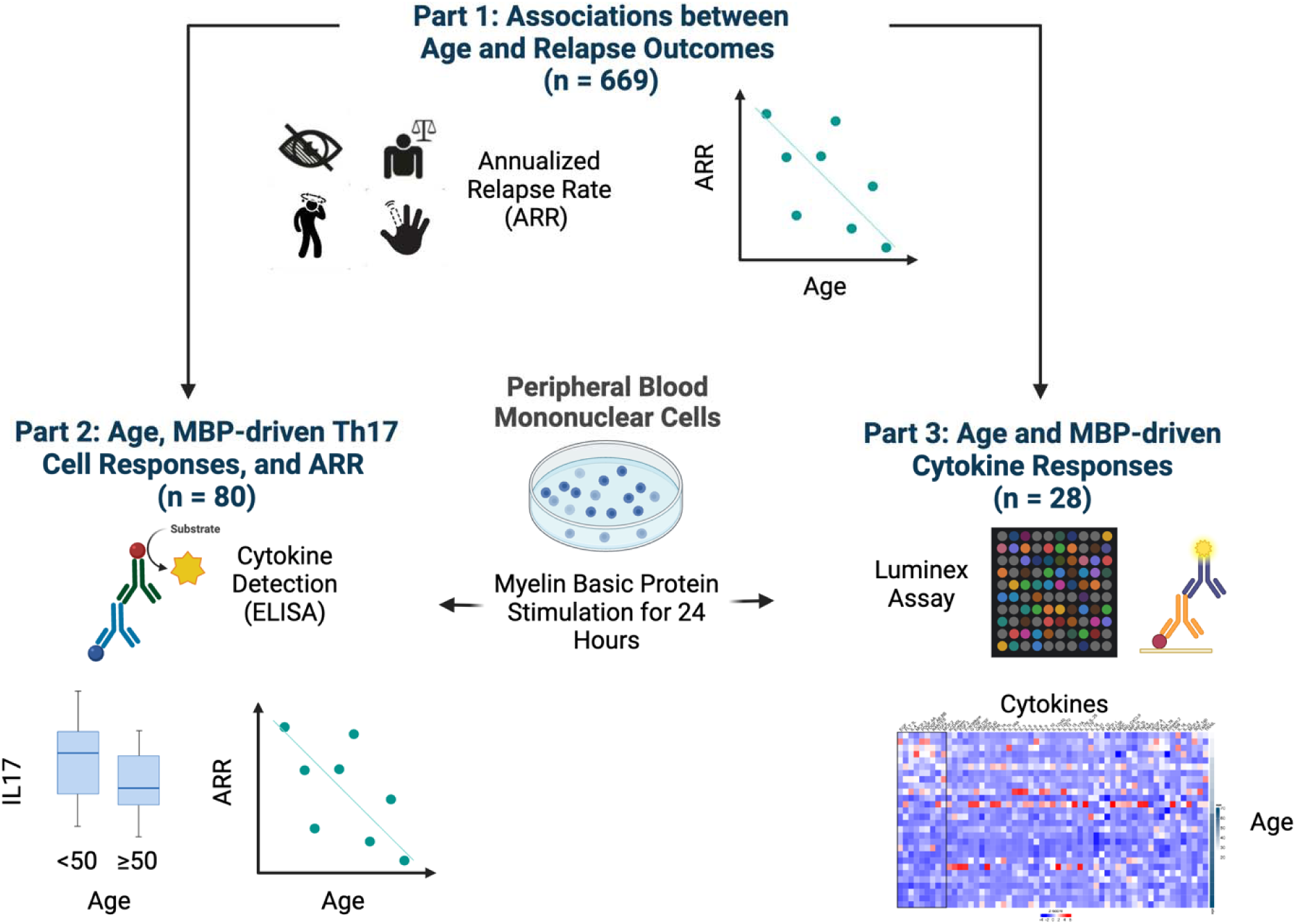
Overall study design.

### Ethics Approval

The institutional review board of the University of Pittsburgh (STUDY19080007) approved the study protocols. All participants provided written informed consent.

### Age as Exposure and Other Covariates

We collected demographic and clinical data, including age (at the time of relapse history annotation [primary cohort] or blood sample collection [Th17 cohort]), sex (designated at birth: men versus women), race/ethnicity (non-Hispanic White versus otherwise, given the relatively low proportion of individuals who are not non-Hispanic White in the clinic population), disease subtype (RRMS versus SPMS), disease duration, and use of disease modifying therapy [DMT] (high-efficacy versus standard-efficacy versus no DMT at the time of relapse annotation [primary cohort] or blood sample collection [Th17 cohort]) through review of the electronic health records (EHR). Disease duration was defined as the time interval between MS diagnosis and the time of relapse history annotation (primary cohort) or blood sample collection (Th17 cohort). For DMT, we categorized natalizumab, ocrelizumab, and rituximab as higher-efficacy, while dimethyl fumarate, fingolimod, glatiramer acetate, interferon beta and teriflunomide as standard-efficacy.

### Relapse Rate as the Clinical Outcome

We annotated relapse events through EHR review. A relapse event could be a clinical and/or radiologic relapse. Clinical relapses are either new or recurrent neurological symptoms (relevant to MS) lasting persistently for 24 hours or longer without fever or infection. Radiological relapses are new T1-enhancing lesion(s) and/or new or enlarging T2 fluid attenuated inversion recovery (FLAIR) hyperintense lesion(s) based on clinical radiology reports of routine brain, orbit, or spinal cord magnetic resonance imaging (MRI) studies. Annualized relapse rate (ARR) is a commonly used MS relapse outcome and computed as: ARR = number of relapses / total person-years. For the primary cohort, we calculated the annualized relapse rate (ARR) in the two years preceding the annotation of relapse history. For the Th17 cohort, we calculated the ARR encompassing the two years preceding and the two years after the blood sample collection.

### MBP Stimulation Assay

From freshly collected whole blood, we isolated peripheral blood mononuclear cells (PBMCs) by Ficoll-plague (Sigma-Aldrich) density gradient centrifugation (400x g) for 30 min at room temperature with no brake. PBMCs were collected, washed twice with PBS at (200x g) for 15 min at 4°C, and then cultured at a density of 2×10^6^ cells per well in 24-well plate of human MBP antigen (50 μg/mL, catalog #M0689, Sigma Aldrich) or heat-killed Candida (HKC) as positive control (0.5 million, strain SC5314, a kind gift from Dr. Sarah Gaffen) for 24 hours.[28] Our prior unpublished preliminary experiments showed more consistent assay results using 24 hours of culture than longer periods of incubation.

### MBP-driven Cytokines

We collected supernatants of cultured PBMCs after 24-hours ex vivo MBP or HKC stimulation and performed enzyme-linked immunosorbent assay (ELISA) using the Ready-SET-Go kit for human IL-17 (catalog #88-7176-88) and IFN-γ (catalog #88-7316-88) (eBioscience) according to the manufacturer’s instructions. We performed ELISA assays of each supernatant sample in duplicates. We first calculated the concentration of IL-17 or IFN-γ in each sample (MBP or HKC-stimulated PBMC) by subtracting background concentration (unstimulated PBMC), and then reported the mean concentrations of the duplicates. In addition, we analyzed a subset of supernatant samples assayed using the Luminex xMAP platform human cytokine/chemokine array (Eve Technologies, Canada).

### Statistical Analysis

To select the covariates for adjustment in downstream analysis, we first performed univariate linear regressions to assess associations between potentially relevant clinical or demographic features (sex, race/ethnicity, disease subtype and DMT efficacy) and the MS relapse outcome (*i.e.*, ARR). We did not include disease duration as a covariate given its high correlation with the exposure of interest (*i.e.,* age), which would have caused over-correction. Next, we assessed the associations between (1) age and ARR, (2) age and (concentration of) MBP-stimulated cytokines, and (3) MBP-stimulated cytokines and ARR, using multi-variate linear regressions accounting for confounders deemed significant in the univariate analyses. Finally, we performed mediation tests to compute the proportion of the association between age and ARR explained by the MBP-driven cytokine responses (Th17/IL-17 and other cytokines). The total effect of the exposure (*i.e.,* age) on the outcome (*i.e*., ARR) encompasses both direct effect (of the exposure on the outcome) and indirect effect mediated through one or more intermediaries (*e.g.*, cytokine).[43] A two-tailed p-value <0.05 was deemed statistically significant. We performed all analyses using R, version 4.0.2.

## Results

### Inverse association between age and relapse rate in pwMS

The study **primary cohort** included the clinical data of 669 adult pwMS (mean age 51.7±12.7 years, 80.7% women, 89.4% non-Hispanic White) who enrolled in a clinic-based MS cohort between 2017 and 2022 (Table 1). Among the 669 participants, univariate analyses showed that only sex, but *not* race/ethnicity, disease subtype, or DMT efficacy, was significantly associated with ARR. Thus, we did not include race/ethnicity, disease subtype or DMT status in multivariable analyses. To reduce over-correction, we also did not include disease duration as a covariate in the analyses because of its strong correlation with age. After adjusting for confounder selected by the univariate analyses (*i.e.*, sex), age was inversely associated with ARR (beta = -0.003, 95% CI [-0.005, -0.002], p<0.001) in the primary cohort (Table 2). In a randomly selected subset of 80 participants that constitute the **Th17 cohort** who shared mostly similar characteristics as the primary cohort (Table 1) and who donated PBMCs for the ex vivo MBP-stimulation assays, the inverse association between age and ARR remained significant after covariate adjustment (beta = -0.016, 95%CI [-0.024, -0.005], p=0.002). Thus, as a crucial sanity check, aging-associated decline in relapse rate in the study cohort is consistent with the existing literature and paves the way for mechanistic explorations.

**Table 1.** Cohort characteristics.

|  | Primary Cohort |  |  | Th17 Cohort |  |  |
| --- | --- | --- | --- | --- | --- | --- |
|  | All<br>(n = 669) | Women<br>(n = 540) | Men<br>(n = 129) | All<br>(n = 80) | Women<br>(n = 56) | Men<br>(n = 24) |
| **Age**<br>**(years, mean ± SD)** | 51.7 (12.7) | 51.9 (12.4) | 50.8 (13.8) | 47.15 (13.3) | 47.3 (13.5) | 46.8 (13.3) |
| **Race / Ethnicity**<br>**(% of cohort)** |  |  |  |  |  |  |
| Non-Hispanic White | 598 (89.4) | 478 (88.5) | 120 (93.0) | 74 (86.0) | 48 (85.7) | 21 (87.5) |
| Other | 71 (10.6) | 62 (11.5) | 9 (7.0) | 12 (14.0) | 8 (14.3) | 3 (12.5) |
| **Disease Duration**<br>**(years, mean ± SD)** | 14.0 (9.6) | 14.3 (9.5) | 12.8 (10.2) | 13.34 (10.15) | 14.3 (10.6) | 12.5 (9.6) |
| **Disease Subtype**<br>**(n, % of cohort)** |  |  |  |  |  |  |
| RRMS | 638 (95.4) | 522 (96.7) | 116 (89.9) | 78 (97.5) | 56 (100.0) | 22 (91.7) |
| SPMS | 31 (4.6) | 18 (3.3) | 13 (10.1) | 2 (2.5) | 0 (0.0) | 2 (8.3) |
| **DMT Treatment**<br>**(n, % of cohort)** |  |  |  |  |  |  |
| No DMT | 185 (27.7) | 154 (28.5) | 31 (24.0) | 16 (20.0) | 13 (23.2) | 3 (12.5) |
| Standard-efficacy DMT | 166 (24.8) | 132 (24.4) | 34 (26.4) | 23 (28.7) | 20 (35.7) | 3 (12.5) |
| High-efficacy DMT | 318 (47.5) | 254 (47.1) | 64 (49.6) | 41 (51.2) | 23 (41.1) | 18 (75.0) |

Abbreviations: *SD*, standard deviation; *RRMS*, relapse-remitting MS; *SPMS*, secondary progressive MS; *DMT*, disease-modifying therapy. (Please refer to Methods for the designation of DMT efficacy categories.)

**Table 2.** Age is inversely associated with annualized relapse rate (ARR) in pwMS.

|  | Primary Cohort<br>(n = 669) |  | Th17 Cohort<br>(n = 80) |  |
| --- | --- | --- | --- | --- |
|  | Coefficient <sup>4</sup><br>(95%CI) | p-value | Coefficient <sup>4</sup><br>(95%CI) | p-value |
| **Unadjusted**<br>**Linear Regression <sup>1</sup>** |  |  |  |  |
| ARR <sup>3</sup> | -0.003<br>(-0.004, -0.002) | < 0.001 | -0.016<br>(-0.017, -0.006) | < 0.001 |
| **Adjusted**<br>**Linear Regression <sup>2</sup>** |  |  |  |  |
| ARR <sup>3</sup> | -0.003<br>(-0.005, -0.002) | < 0.001 | -0.016<br>(-0.024, -0.005) | 0.002 |

Note:
1. Unadjusted linear regression: ARR ~ age. 2. Adjusted linear regression: ARR ~ age + sex. Please note that the multivariable analyses only adjusted for confounder deemed significant in the univariate analyses (*i.e.*, the feature selection step). 3. Annualized relapse rate (ARR) definition: In the primary cohort, ARR was calculated in the 2 years preceding the relapse history annotation date. In the Th17 cohort, ARR was calculated within ± 2 years of sample collection. 4. Coefficient for age. Abbreviations: *CI*, confidence interval

### Inverse association between age and MBP-driven Th17 responses in women with MS

To assess the association between age and MBP-driven Th17/IL-17 response, we considered age as either a continuous variable or a binary variable using ≥50 versus <50 years as the threshold. The binary age threshold is based on prior clinical studies examining the age-related decline in MS relapse rate.[5,6] In the full Th17 cohort, age was *not* significantly associated with MBP-driven IL-17 expression (Figure 2A and D). Interestingly, there was a significant inverse association between age and MBP-driven IL-17 response in women (age as a continuous variable: beta = -0.268, 95%CI [-0.519, -0.018], p=0.04; age as a binary variable: p=0.031) but not in men (Fig 2B and 2E versus Fig 2C and 2F). When assessing the association between the MBP-driven IL-17 response and ARR, we found significant positive correlations in women (beta = 0.007, 95%CI [0.002, 0.012], p=0.01) but not in men (Figure 3). While the Th17 cohort included fewer men than women (reflecting the typical ratio of men versus women in MS), the difference in sample size alone seems insufficient to explain the null finding for men given also the null finding in the full Th17 cohort that included *both* men and women. On a related note, the available subgroup sample size as stratified by DMT status at sample collection would be insufficient for comparison across DMTs (and the univariate analysis did not select DMT as a feature associated with relapse).

**Figure 2.**
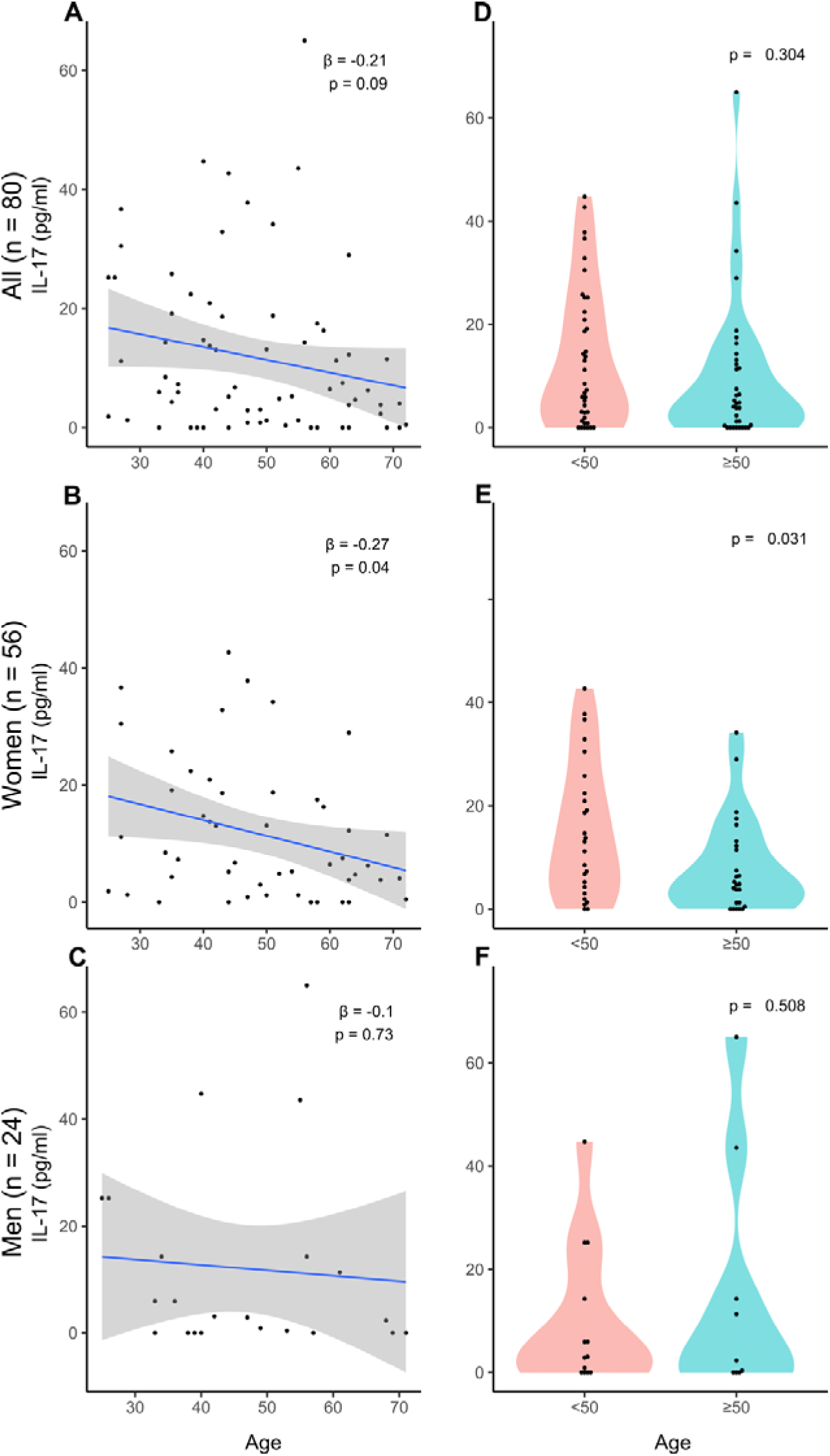
MBP-driven Th17 response was inversely associated with age in women but not in men with MS. PBMCs from pwMS were stimulated ex vivo with MBP for 24 hours. IL-17 concentration in the supernatant was measured using ELISA and plotted against age as a continuous variable (A-C) and age as a binary variable (D-F) using the age threshold of ≥50 years versus <50 years.

**Figure 3.**
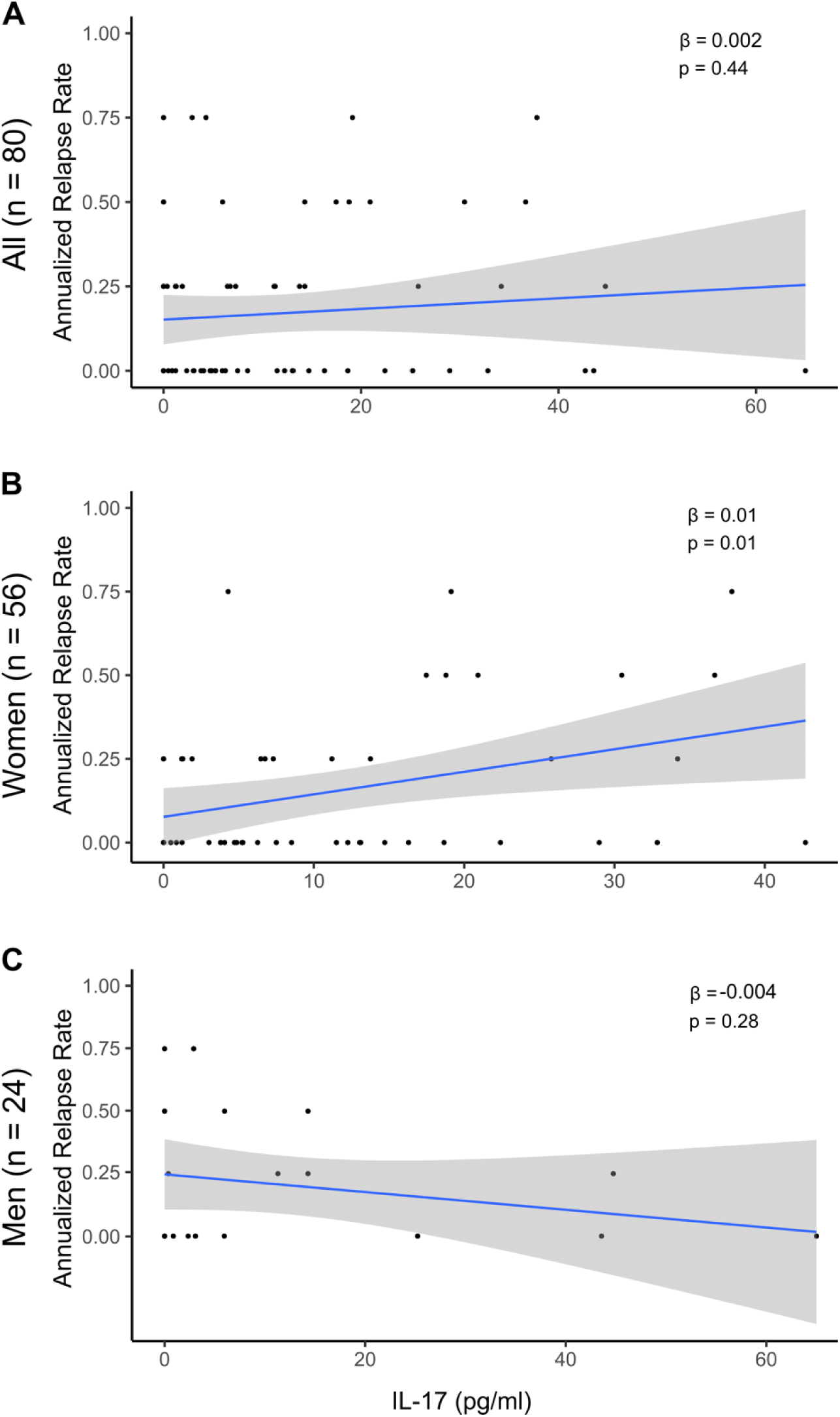
MBP-driven Th17 response was inversely associated with relapse rate in women but not in men with MS. PBMCs from pwMS were stimulated ex vivo with MBP for 24 hours. IL-17 concentrations were plotted against annualized relapse rate (ARR) within ± 2 years of sample collection.

To quantify the extent to which MBP-driven IL-17 response explains the inverse association between age and ARR, we performed mediation analysis (Table 3). Specifically, MBP-driven IL-17 response mediated 24.6% of the effect of older age on lower ARR in women versus 15.3% in men. Overall, these findings support further investigation of the sex differences in MBP-driven peripheral Th17 response in larger cohorts.

**Table 3.**
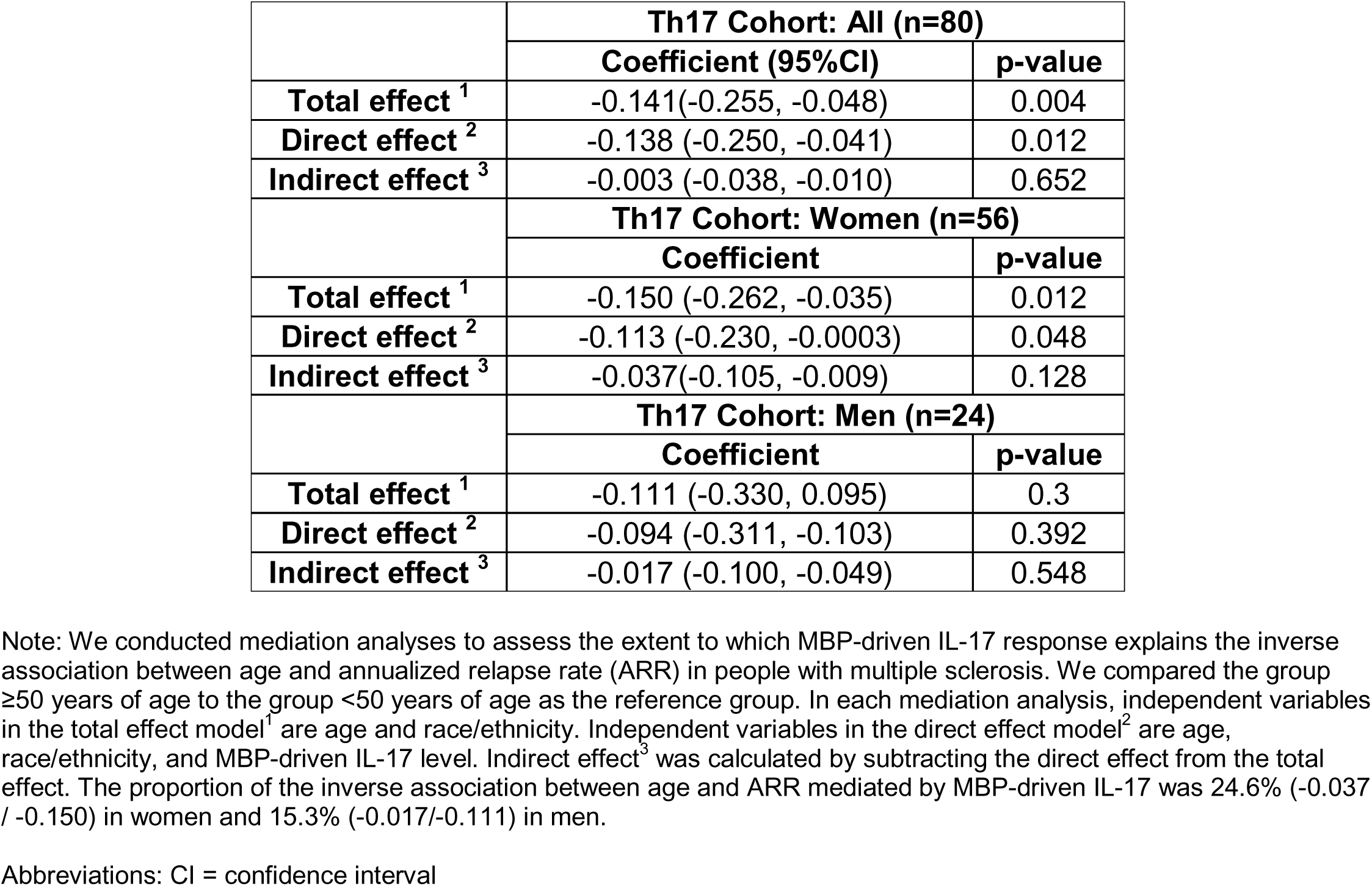
MBP-driven Th17 response as a mediator of the inverse association between age and MS relapse.

IFN-γ is produced by Th1 cells and Th17 cells. We similarly measured MBP-driven IFN-γ responses and found no significant association between age and MBP-driven IFN-γ response or MBP-driven IFN-γ response and ARR in the Th17 cohort (all, women, men) (Supplementary Figure 1). Further, age was not significantly associated with HKC-driven IL-17 or IFN-γ responses (Supplementary Figure 2), suggesting that the aging-dependent decline in MBP-driven IL-17 responses might be a MS-specific finding.

### Age and other MBP-driven cytokine responses: Exploratory Analyses

Given that other cytokines beyond IL-17 might also change with aging in pwMS, we performed *exploratory* analysis of additional MBP-driven cytokine responses in 28 samples from the Th17 cohort (women=20, men=8) using the Luminex xMAP platform human cytokine/chemokine array. We did not stratify the samples by sex in these exploratory analyses due to the more modest sample size. We assayed 57 MBP-driven cytokine responses across the 28 samples and organized the findings in order of ascending age (Figure 4A). Seven cytokines showed significant associations with age when evaluating age as either a continuous or a binary variable (Figure 4B, Figure 5, Supplementary Table 1). In this pilot data set, age as a continuous variable was positively associated with MDC (beta = 9.649, 95%CI [4.808, 14.489], p<0.001) and marginally with Fractalkine (beta = 0.238, 95%CI [0.000, 0.478], p=0.051), but inversely associated with IL-4 (beta = -0.004, 95%CI [-0.009, -3.943], p=0.048), MCP-2 (beta = -12.350, 95%CI [-22.406, -2.295], p=0.018), MCP-3 (beta = -24.218, 95%CI [-41.476, -6.961], p=0.008), PDGF-AA (beta = -0.851, 95%CI [-1.504, -0.197], p=0.013), and PDGF-AB/BB (beta = -7.154, 95%CI [-11.232, -3.075], p=0.001) (Figure 5). There were similarly significant associations when using age as a binary variable. We consider these associations all as *marginally significant* since none survived multiple testing correction for false discovery rate (Supplementary Table 1). We also explored the extent to which these seven marginally significant MBP-driven cytokine responses mediated the association between older age and lower ARR. Two cytokines mediated the effect of age on ARR in similar proportions: MCP-2 (4.15%), MDC (4.53%). Interestingly, other cytokines (Fractalkine, IL-4, MCP-3, PDGF-AA, and PDGF-AB/BB) might negate the effect of age on ARR with the proportion of cytokine-negated effect ranging from 0.75% (PDGF-AB/BB) to 16.98% (IL-4) (see details in Supplementary Table 2).

**Figure 4.**
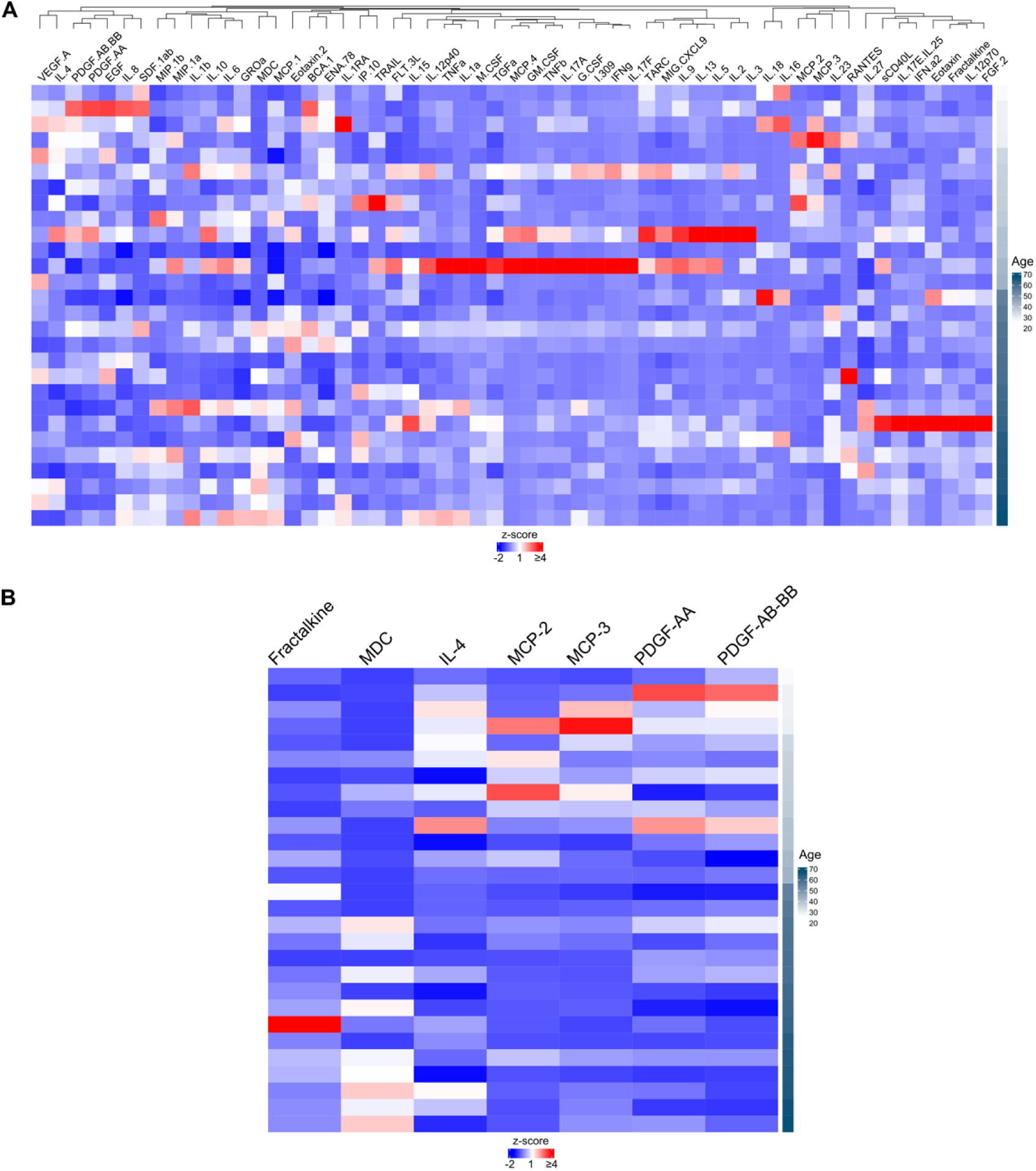
Levels of additional MBP-stimulated cytokines in relation to age. PBMCs from pwMS were stimulated ex vivo with MBP for 24 hours. In exploratory analyses, concentrations of 57 additional cytokines in the supernatant were measured using the Luminex xMAP platform human cytokine/chemokine array. Patient-level Z-score transformed levels of all cytokines are shown (A). Seven cytokines with marginally significant association with age are highlighted (B). Each row represents a unique participant in ascending order of age.

**Figure 5.**
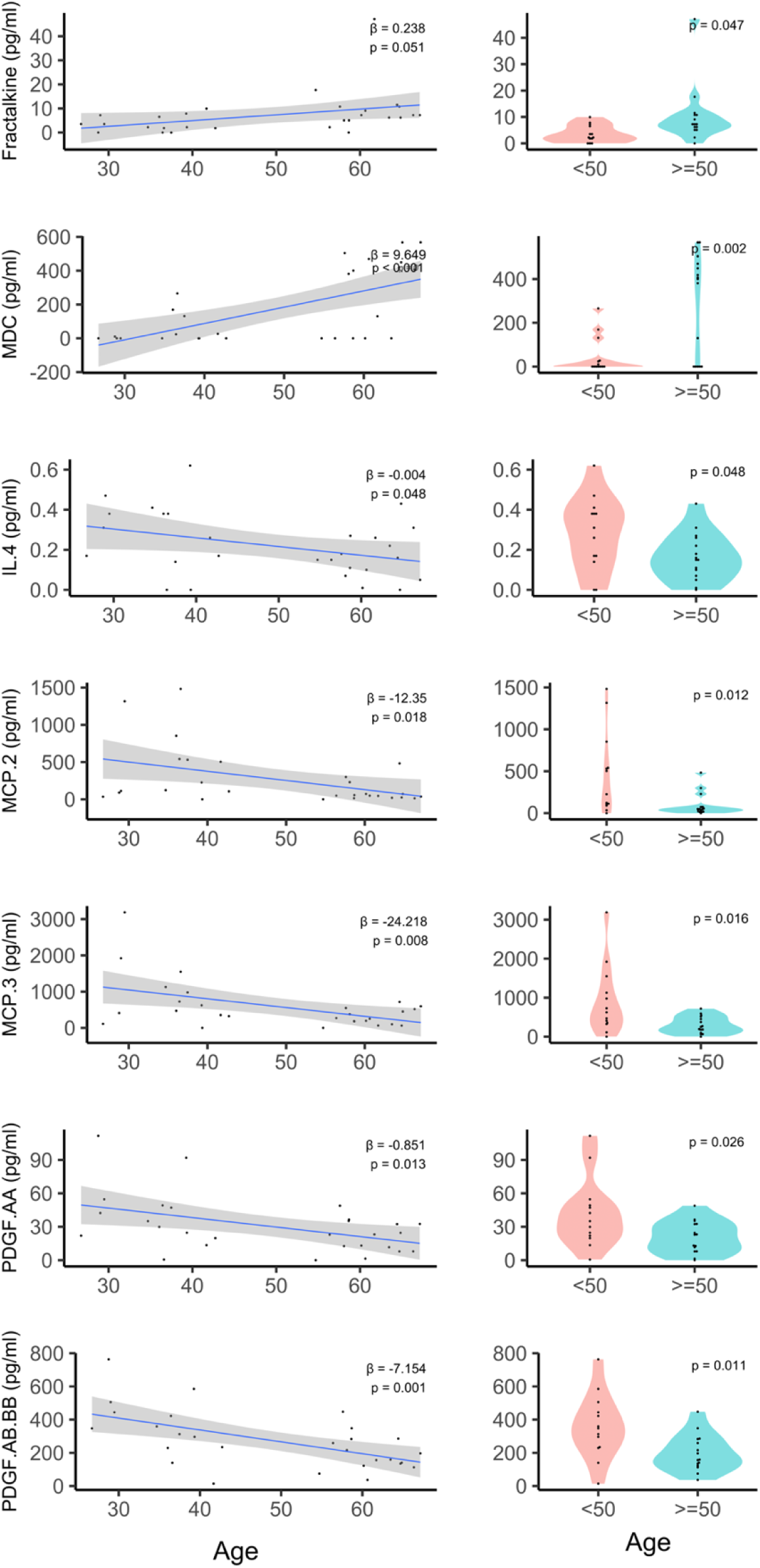
Exploratory associations between age and the additional MBP-stimulated cytokines. The levels of the seven cytokines (from Figure 4B) in relation to age as continuous variable are shown in scatter plots (left panels) and age as a binary variable using the age threshold of ≥50 years versus <50 years are shown in violin plots (right panels).

## Discussion

In this clinic-based cohort study, we report a potential mechanism to explain the clinical observation of the declining relapse rate (*i.e.,* ARR) with increasing age in pwMS that has been the basis for randomized clinical trials of DMT discontinuation in older pwMS. Specially, MBP-driven IL-17 response, but not MBP-stimulated IFN-γ response (or HKC-stimulated IL-17 or IFN-γ response), declined with increasing age in women but not men with MS. MBP-driven IL-17 response partially mediated the effect of older age on lower ARR to a greater extent in women (24.7%) than men (15.3%). In exploratory analyses of other MBP-driven cytokines beyond IL-17, older pwMS (50 years and older) exhibited lower concentrations of IL-4, MCP-2, MCP-3, PDGF-AA, PDGF-AB/BB and higher concentrations Fractalkine and MDC than younger pwMS (below 50 years of age).

This study has several strengths. First, this is the first study to our knowledge that examined the potential peripheral immune mechanisms of the declining inflammatory disease activity with aging in pwMS using the practical experimental paradigm of patient-derived PBMCs stimulated ex vivo with MBP, an abundant CNS myelin protein and a putative autoantigen. This well-established approach enables the investigation of potential changes in MS-relevant autoimmune inflammatory response and associated biomarkers of clinical relevance in older pwMS. Second, this study included crucial controls (*e.g.,* MBP-stimulated IFN-γ response, HKC-stimulated IL-17 and IFN-γ response) that support the MS-specific finding of aging-dependent decline in MBP-driven Th17/IL-17 response (versus non-specific T cell response to general stimulation). Third, we performed mediation analyses to quantify the extent to which a given MBP-driven cytokine response mediates the aging-related decline in inflammatory disease activity in pwMS, enabling comparison between men and women as well as across different cytokines and increasing the interpretability of the findings. Fourth, this study examined the sex difference in potential immune mechanisms underlying aging in MS. Although sex difference in MS susceptibility is well recognized, little is known regarding the sex difference in peripheral immune responses in older pwMS. We found that the extent to which MBP-driven Th17 response mediated aging-related decline in MS inflammatory disease activity (*i.e.,* relapse rate) was greater in women than men. Finally, we explored a broader group of MBP-driven cytokine or chemokine responses beyond Th17/IL-17 that may additionally mediate or negate inflammatory disease activity in aging pwMS. These findings collectively generate intriguing hypotheses for future studies of the mechanisms of aging in MS and age- and sex-specific biomarkers of the MS disease course.

The *major* study findings of the sex-difference (*i.e.,* significant aging-associated decline in MBP-driven Th17 response in women but not men with MS, a greater extent to which MBP-driven Th17 response-mediated aging-associated decline in MS inflammatory disease activity in women than in men) were unlikely to be solely attributable to the differential sample size between men and women. While women who represent a subset of the Th17 cohort exhibited significantly inverse association between age and MBP-driven Th17 response, age was *not* associated with MBP-driven Th17 response in the *full* Th17 cohort that included both men and women, highlighting the strong effect size of age in women. Crucially, several other lines of evidence in the literature support our results. Peripheral immune drivers of MS inflammatory disease activity are postulated to differ by sex, which may support the clinical observation that MS diagnosis is more common in women than men while men with MS is prone to develop progressive disease and disability progression than women.[44] At least two concurrent pathogenic processes across the MS disease stages are potentially relevant to aging in MS.[45,46] First, acute inflammatory demyelination and relapse events are underscored by the migration of peripheral CD8^+^ and CD4^+^ T cells and CD20^+^ B cells into the CNS. Second, progressive neurodegeneration and disability accrual are driven by microglia and macrophage activation, oxidative injury, and mitochondrial damage that can begin concurrently in the early stages of the disease and involve the gradual accumulation of late-differentiated T and B cells in the meninges and periventricular spaces. The development of these two processes in MS seem to have sex differences. A single-cell transcriptomic analysis examining the effects of sex and age on the peripheral immune cell landscape in the general human population reported higher percentage of plasma cells and over-representation of gene expression pathways relevant to the adaptive immune function (B and T cell activation) in women versus higher percentage of NK cells and higher expression of pro-inflammatory genes in men. Importantly, aging further amplified the observed sex differences.[47] Dysregulation of T-cell subtypes occurs with normal aging in the broader population, manifesting paradoxically as an aging-dependent increase in Th17/Treg ratio at baseline but reduced Th17/Treg ratio after stimulation (by phytohaemagglutinin, a potent stimulator of IL-17 response in PBMCs though not necessarily specific for MS).[19] Menopause in women is a potential aging-related inflection point (around 40-50 years of age for most women) in immune alteration, accompanied by an increase in Th17/Treg ratio.[48] Given the current understanding that adaptive immunity (*e.g.,* B and T cell activation) is generally more robust in women than men while aging-dependent Th17/Treg dysregulation coincides with the perimenopausal period (a key milestone in female aging) and exposure to non-MS specific stimulant of IL-17 paradoxically reduces the aging-dependent Th17/Treg ratio, it is conceivable that the aging-dependent decline in MS relapse rate may be driven by the diminishing MS-specific (*e.g.,* MBP-driven) IL-17 responses to a greater extent in women than men, as reported in this study. Overall, our study findings contribute to the broader literature on sex-difference in the aging of human immune systems.[49]

The additional *exploratory* study findings of the other MBP-driven cytokine profile beyond Th17 response in older pwMS (50 years and older) (*i.e.,* significantly lower concentrations of IL-4, MCP-2, MCP-3, PDGF-AA, PDGF-AB-BB, significantly higher concentrations Fractalkine and MDC secreted by MBP-stimulated PBMCs) when compared to pwMS below 50 years of age generate testable hypotheses for future studies. We briefly summarize the prior literature regarding the seven cytokine / chemokines in MS, but *no prior study* to our knowledge examined aging-associated changes in these molecules. IL-4 levels are elevated in blood and cerebrospinal fluid (CSF) samples from pwMS.[50–52] The MCP family of chemokines are expressed in MS lesions in the CNS and may contribute to lesion formation.[53] PDGF isoforms regulate long-term synaptic protection and adaptive plasticity and may promote functional recovery in RRMS, while serum levels of PDGF are lower in pwMS than controls.[54–56] Fractalkine (*i.e.,* CX3CL1) recruits subsets of CD4 T cells across the blood-brain barrier into the CNS during early MS, though its site of action seems peripheral.[57–59] MDC (*i.e.,* CCL22) levels are lower in the serum and higher in the CSF of female (but not male) pwMS than female controls.[56,60–62] In our study, lower levels IL-4, MCP-2, MCP-3 and higher level of MDC (by MBP-stimulated PBMCs) in older pwMS were potentially consistent with the observation of reduced IL-17 level and the lower relapse rate with aging. Further, we speculate that the increased fractalkine versus reduced PDGF production could be associated with the long-term loss of function in the aging MS brain. To better examine the effect of cytokines in the CNS, future studies need to go beyond the MBP-stimulated PBMC paradigm. Nevertheless, the current study findings suggest further investigation of potential biomarkers of aging-associated switch in the drivers of MS pathology from predominantly pro-inflammatory mechanisms to predominantly neurodegenerative mechanisms.

The main study limitations pertain to the sex imbalance in sample size, low racial and ethnic diversity, a single center cohort and cross-sectional data set. Given that the study population drew from a real-world clinic-based cohort of pwMS in the Western Pennsylvania region of the United States, we are limited by the available research participants who are over-represented by women and non-Hispanic White individuals, largely reflecting the broader clinic population. Further, subgroup analyses based on DMT did not seem justifiable because the pre-planned univariate analysis (for feature selection) in this study did not select DMT status for adjustment in downstream multivariable analysis (*e.g.,* age and ARR). Consideration for past DMT history beyond DMT status at relapse annotation (or sample collection) may be necessary for future studies of aging-associated relapse rate. Finally, the study sample size was insufficient to assess the role of MBP-Th17 response in the conversion from RRMS to SPMS. Overall, future studies that include a larger sample size, greater proportion of men and historically under-represented racial and ethnic groups, external samples and data sources as well as a prospective study design will be crucial to validate the current findings and enhance robustness. Using the same experimental paradigm, future studies could also assess the potential role of menopause in aging-associated decline in MBP-Th17 response in women, the attributable risk of MBP-Th17 response in the conversion from RRMS to SPMS, and the extent to which different DMT classes may interact with aging-associated decline in MB-Th17 response.

## Conclusion

This study highlights the potentially important role of MBP-driven Th17/IL-17 response and additional peripheral immune cytokine mediators in aging-dependent decline in MS inflammatory disease activity, particular in women with MS.

## Data Availability

Code for analysis and figures is available at <https://github.com/xialab2016/AgeandTh17.git >. De-identified data are available upon request to the corresponding author and with permission from the participating institution.

## Acknowledgment

We appreciate the contribution of research participants and clinicians at the UPMC MS Center.

## Data Sharing

Code for analysis and figures is available at <https://github.com/xialab2016/AgeandTh17.git>. De-identified data are available upon request to the corresponding author and with permission from the participating institutions.

