## Supplementary Figures and Tables for "Aging-dependent Change in Th17 and Cytokine Response in Multiple Sclerosis"

**Supplementary Table 1.** MBP-stimulated cytokine / chemokine responses between binary age groups.

| Molecule Abbreviation | Molecule Full Name | Age <50 (n=13) | Age ≥50 (n=15) | Unadjusted p-value | FDR |
|---|---|---|---|---|---|
| | | mean (SD), pg/ml | mean (SD), pg/ml | | |
| BCA-1 | B cell-attracting chemokine-1 | 26.18 (16.65) | 19.61 (12.87) | 0.25 | 0.55 |
| EGF | epidermal growth factor | 7.04 (6.48) | 4.62 (4.81) | 0.268 | 0.57 |
| ENA-78 | epithelial-derived neutrophil-activating peptide 78 | 7858.54 (3051.66) | 6029.08 (3872.69) | 0.182 | 0.55 |
| Eotaxin | eotaxin | 0.20 (0.38) | 0.86 (1.55) | 0.149 | 0.53 |
| Eotaxin.2 | eotaxin-2 | 1058.22 (1562.06) | 2167.34 (2144.98) | 0.135 | 0.53 |
| FGF-2 | fibroblast growth factor 2 | 2.52 (2.13) | 7.53 (9.85) | 0.084 | 0.53 |
| FLT-3L | fms-like tyrosine kinase receptor 3 ligand | 0.39 (0.41) | 0.28 (0.20) | 0.398 | 0.6 |
| Fractalkine | fractalkine | 3.59 (3.27) | 10.22 (11.02) | 0.047 | 0.39 |
| G.CSF | granulocyte colony stimulating factor | 7.63 (13.39) | 3.59 (5.61) | 0.295 | 0.6 |
| GM-CSF | granulocyte-macrophage colony stimulating factor | 29.32 (62.32) | 4.13 (8.08) | 0.132 | 0.53 |
| GRO-a | growth related oncogene-alpha | 1433.48 (721.58) | 1143.09 (797.93) | 0.325 | 0.6 |
| I-309 | cytokine I-309 | 17.74 (38.15) | 9.80 (8.77) | 0.44 | 0.63 |
| IFN-a2 | interferon alpha 2 | 2.33 (1.82) | 3.42 (5.16) | 0.477 | 0.66 |
| IFN.γ | interferon gamma | 8.23 (16.50) | 1.27 (1.70) | 0.115 | 0.53 |
| IL-1a | interleukin-1 alpha | 2.52 (7.19) | 2.90 (4.15) | 0.86 | 0.98 |
| IL-1b | interleukin-1 beta | 13.09 (17.89) | 17.92 (23.61) | 0.552 | 0.72 |
| IL-1RA | interleukin-1 receptor antagonist | 37.84 (89.09) | 40.13 (47.67) | 0.932 | 0.98 |
| IL-2 | interleukin-2 | 12.41 (24.20) | 6.72 (8.14) | 0.398 | 0.6 |
| IL-3 | interleukin-3 | 2.09 (4.69) | 1.21 (1.46) | 0.5 | 0.66 |
| IL-4 | interleukin-4 | 0.28 (0.18) | 0.16 (0.12) | 0.048 | 0.39 |
| IL-5 | interleukin-5 | 7.38 (15.77) | 1.81 (3.48) | 0.194 | 0.55 |
| IL-6 | interleukin-6 | 686.48 (593.29) | 462.24 (549.82) | 0.309 | 0.6 |
| IL-8 | interleukin-8 | 6508.76 (3969.31) | 5944.49 (3105.28) | 0.677 | 0.84 |
| IL-9 | interleukin-9 | 2.62 (4.11) | 1.11 (1.08) | 0.18 | 0.55 |
| IL-10 | interleukin-10 | 51.96 (46.65) | 38.82 (30.34) | 0.379 | 0.6 |
| IL-12p40 | interleukin-12 p40 subunit | 9.81 (12.40) | 9.85 (9.28) | 0.992 | 0.99 |
| IL-12p70 | interleukin-12 p70 subunit | 0.08 (0.13) | 0.30 (0.50) | 0.14 | 0.53 |
| IL-13 | interleukin-13 | 16.96 (29.08) | 7.14 (7.61) | 0.218 | 0.55 |
| IL-15 | interleukin-15 | 0.42 (0.50) | 0.61 (0.66) | 0.413 | 0.6 |
| IL-16 | interleukin-16 | 33.97 (73.49) | 21.02 (52.42) | 0.592 | 0.75 |
| IL-17A | interleukin-17A | 9.09 (16.55) | 2.60 (2.82) | 0.146 | 0.53 |
| IL-17E (IL-25) | interleukin-17A (also known as interleukin-25) | 0.54 (0.44) | 1.03 (1.28) | 0.205 | 0.55 |

| | | | | | |
|---|---|---|---|---|---|
| IL-17F | interleukin-17F | 34.43 (86.32) | 3.55 (5.16) | 0.177 | 0.55 |
| IL-18 | interleukin-18 | 0.61 (1.09) | 0.66 (1.44) | 0.92 | 0.98 |
| IL-23 | interleukin-23 | 7.60 (9.28) | 6.77 (7.98) | 0.803 | 0.97 |
| IL-27 | interleukin-27 | 14.62 (10.14) | 19.18 (17.02) | 0.407 | 0.6 |
| IP-10 | Interferon gamma-induced protein 10 | 1725.41 (1963.95) | 1836.38 (2045.87) | 0.885 | 0.98 |
| MCP-1 | monocyte chemoattractant protein 1 | 1170.04 (780.66) | 1416.12 (730.53) | 0.397 | 0.6 |
| MCP-2 | monocyte chemoattractant protein 2 | 455.48 (490.72) | 97.93 (135.06) | 0.012 | 0.23 |
| MCP-3 | monocyte chemoattractant protein 3 | 906.47 (883.40) | 300.90 (220.80) | 0.016 | 0.23 |
| MCP-4 | monocyte chemoattractant protein 4 | 140.84 (253.65) | 19.65 (46.88) | 0.08 | 0.53 |
| M-CSF | macrophage colony-stimulating factor | 4.22 (11.13) | 6.31 (3.23) | 0.491 | 0.66 |
| MDC | macrophage-derived chemokine | 48.22 (85.43) | 286.27 (232.03) | 0.002 | 0.11 |
| MIG (CXCL9) | monokine induced by gamma (also known as C-X-C motif chemokine ligand 9) | 1741.65 (2011.68) | 1170.79 (1014.53) | 0.342 | 0.6 |
| MIP-1a | macrophage inflammatory protein 1 alpha | 34.89 (49.45) | 34.37 (49.23) | 0.978 | 0.99 |
| MIP-1b | macrophage inflammatory protein 1 beta | 181.07 (172.08) | 190.56 (152.31) | 0.878 | 0.98 |
| PDGF-AA | platelet-derived growth factor AA | 41.74 (30.94) | 20.72 (14.36) | 0.026 | 0.3 |
| PDGF-AB/BB | platelet-derived growth factor AB/BB | 357.92 (194.32) | 198.19 (109.85) | 0.011 | 0.23 |
| RANTES | regulated on activation, normal T cell expressed and secreted (also known as C-C motif chemokine ligand 5 | 18.57 (49.27) | 57.93 (107.24) | 0.235 | 0.55 |
| sCD40L | soluble CD40 ligand | 2.02 (3.17) | 3.22 (4.17) | 0.408 | 0.6 |
| SDF-1a+b | stromal cell-derived factor 1alpha + beta | 17.75 (25.39) | 17.60 (18.65) | 0.986 | 0.99 |
| TARC | thymus and activation-regulated chemokine | 25.29 (34.49) | 10.10 (13.04) | 0.125 | 0.53 |
| TGFa | transforming growth factor alpha | 1.40 (1.60) | 1.33 (0.80) | 0.886 | 0.98 |
| TNFa | tumor necrosis factor alpha | 59.83 (109.28) | 63.14 (62.58) | 0.921 | 0.98 |
| TNFb | tumor necrosis factor beta | 5.36 (7.87) | 2.86 (1.77) | 0.24 | 0.55 |
| TRAIL | tumor necrosis factor-related apoptosis-inducing ligand | 0.88 (1.60) | 0.35 (0.47) | 0.229 | 0.55 |
| VEGF-A | vascular endothelial growth factor A | 49.04 (48.17) | 35.23 (36.30) | 0.396 | 0.6 |

Note: The molecules highlighted in red reached marginal level of significance for association with binary age, but none survived multiple testing correction.

Abbreviation: *FDR,* false discovery rate accounting for multiple testing.

**Supplementary Table 2.** MBP-driven response of selected molecules (see Supplementary Table 1) as mediators of the association between age and annualized relapse rate.

| n=28 (women=20, men=8) | | |
|---|---|---|
| **Fractalkine** | **Coefficient (95%CI)** | **p-value** |
| **Total effect** | -0.265 (-0.410, -0.125) | <0.001 |
| **Direct effect** | -0.270 (-0.428, -0.080) | 0.004 |
| **Indirect effect** | 0.005 (-0.136, -0.0446) | 0.956 |
| **MDC** | | |
| **Total effect** | -0.265 (-0.410, -0.125) | <0.001 |
| **Direct effect** | -0.254(-0.457, -0.059) | 0.016 |
| **Indirect effect** | -0.012 (-0.113, -0.090) | 0.804 |
| **IL-4** | | |
| **Total effect** | -0.265 (-0.410, -0.125) | <0.001 |
| **Direct effect** | -0.310 (-0.460, -0.153) | <0.001 |
| **Indirect effect** | 0.045 (-0.037, 0.134) | 0.276 |
| **MCP-2** | | |
| **Total effect** | -0.265 (-0.410, -0.125) | <0.001 |
| **Direct effect** | -0.255 (-0.388, -0.091) | 0.004 |
| **Indirect effect** | -0.011(-0.126, -0.098) | 0.956 |
| **MCP-3** | | |
| **Total effect** | -0.265 (-0.410, -0.125) | <0.001 |
| **Direct effect** | -0.278 (-0.450, -0.108) | <0.001 |
| **Indirect effect** | 0.013 (-0.076, 0.078) | 0.604 |
| **PDGF-AA** | | |
| **Total effect** | -0.265 (-0.410, -0.125) | <0.001 |
| **Direct effect** | -0.286 (-0.462, -0114) | <0.001 |
| **Indirect effect** | 0.020 (-0.051, 0.113) | 0.616 |
| **PDGF-AB/BB** | | |
| **Total effect** | -0.265 (-0.410, -0.125) | <0.001 |
| **Direct effect** | -0.267 (-0.448, -0.099) | 0.008 |
| **Indirect effect** | 0.002 (-0.075, 0.103) | 0.984 |

Note: We conducted mediation analyses to assess the extent to which MBP-driven cytokine or chemokine response (selected from the list in Supplementary Table 1) explains the association between age and annualized relapse rate in people with multiple sclerosis. We compared the group ≥50 years of age to the group <50 years of age as the reference group. In each mediation analysis, independent variables in the total effect model are age and race/ethnicity. Independent variables in the direct effect model are age, race/ethnicity and MBP-driven cytokine or chemokine level. Indirect effect was calculated by subtracting direct effect from total effect. We consider a negation when the indirect effect is in the opposite direction as the direct or total effect.

Abbreviations: CI, confidence interval

**Supplementary Figure 1. MBP-driven IFN-γ response was not associated with age** as either a continuous variable (A-C) or binary variable (D-F).

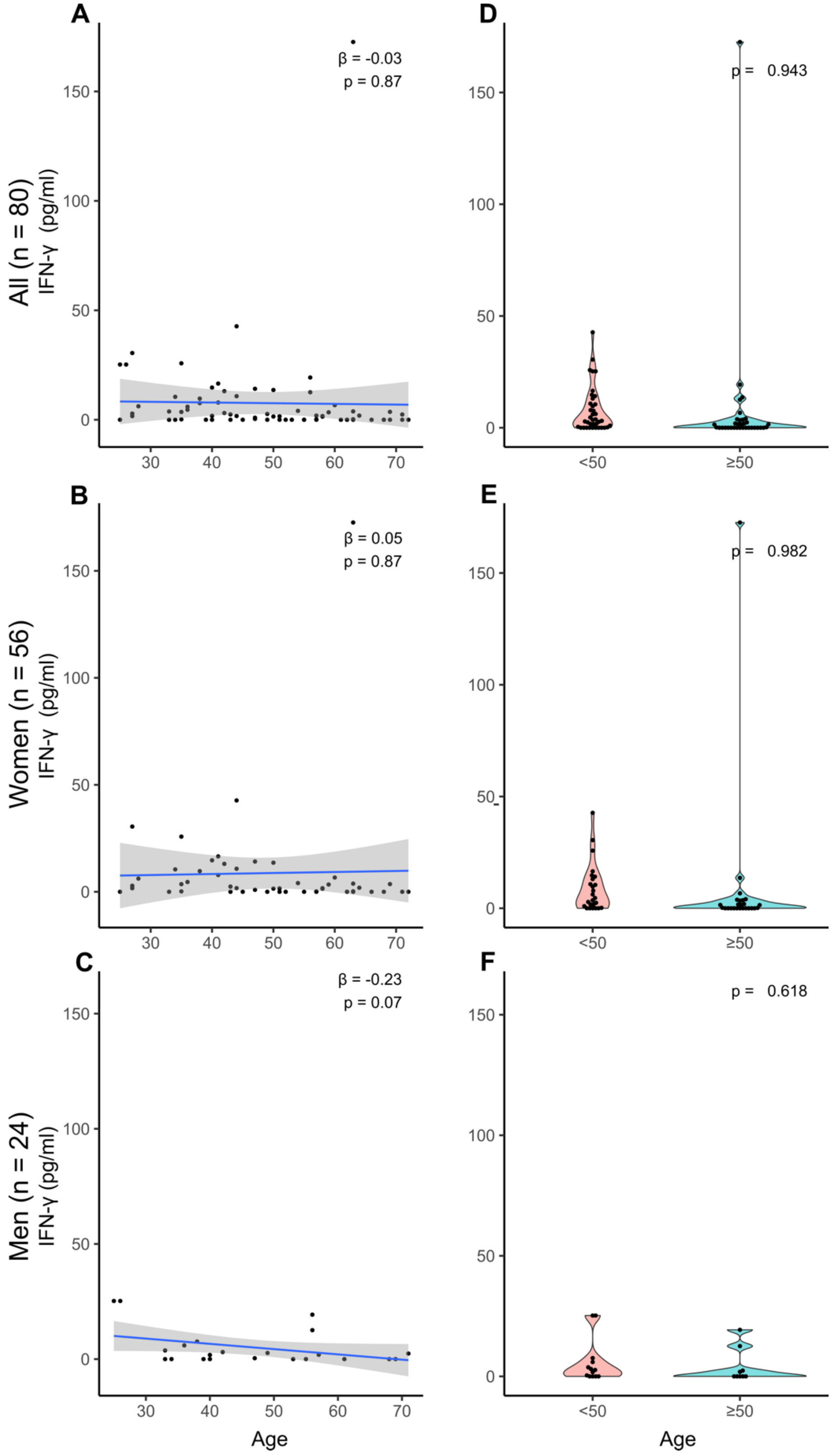

**Supplementary Figure 2. HKC-driven IL-17 response was not associated with age** either as a continuous variable (A-C) or binary variable (D-F).

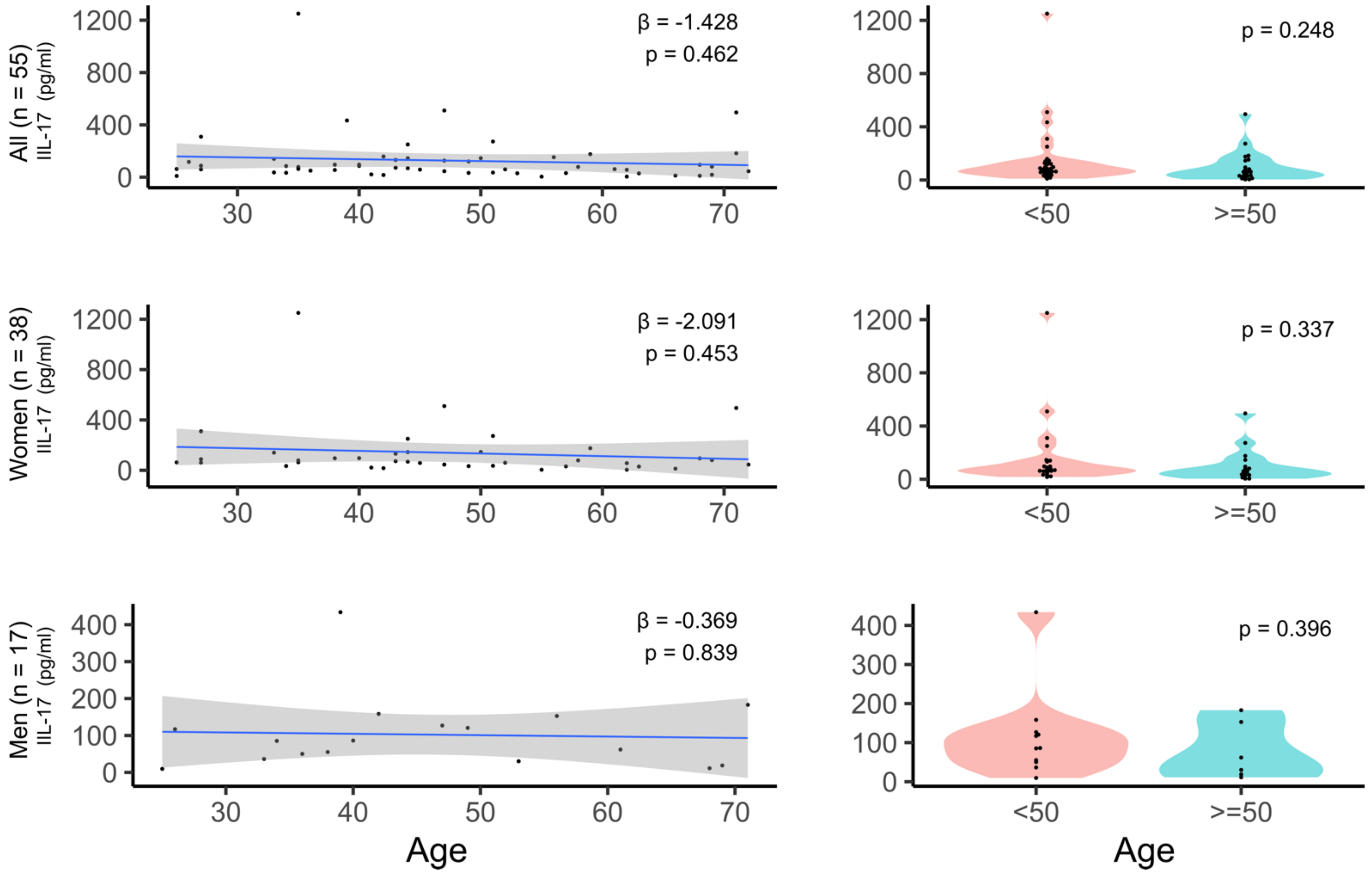

**Supplementary Figure 3. HKC-driven IFN-γ response was not associated with age** either as a continuous variable (A-C) or binary variable (D-F).

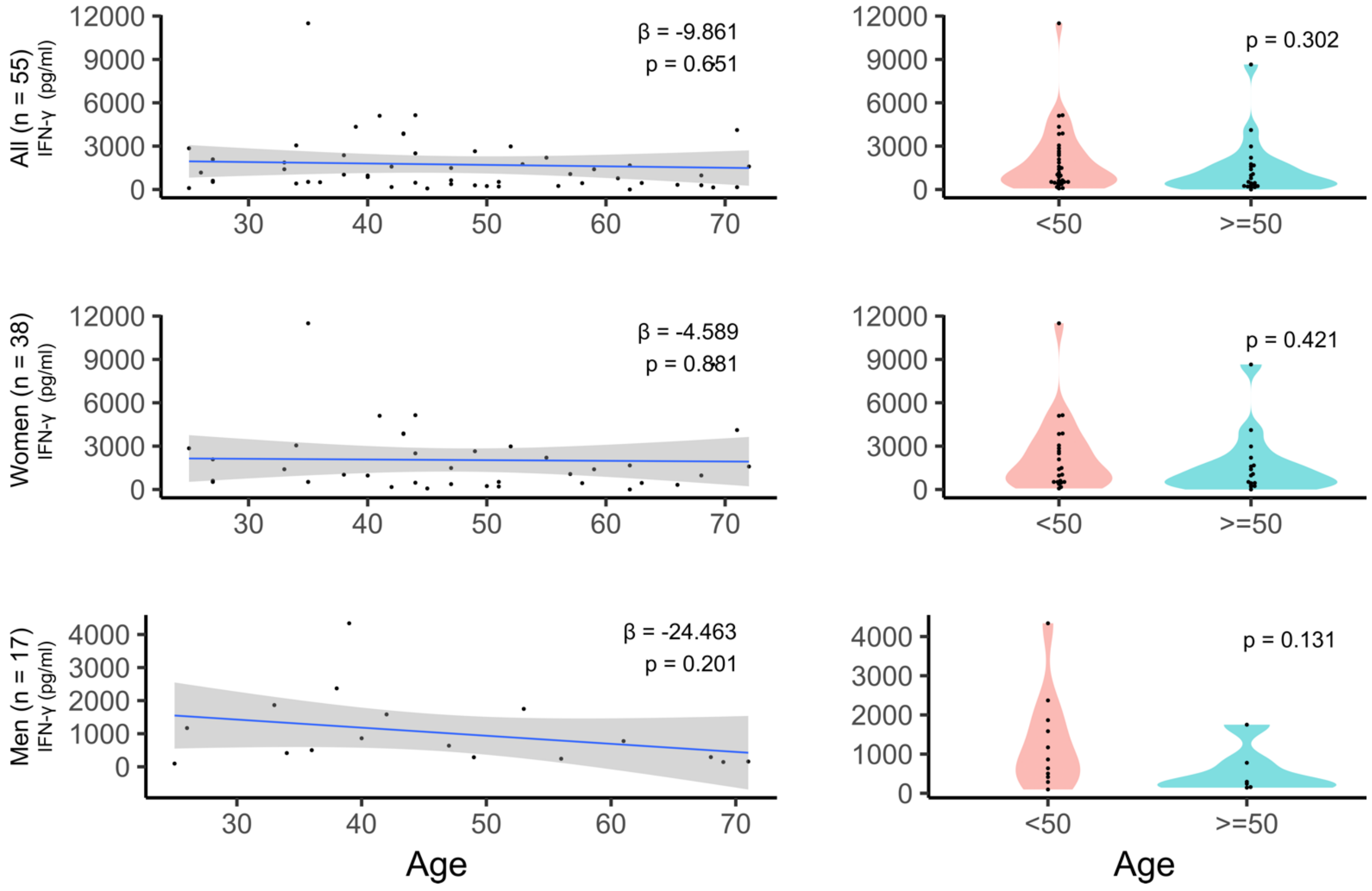
